# Maternal Mortality and Last Mile Challenges: Improved Perspective for Leapfrogging

**DOI:** 10.1101/2023.11.08.23298278

**Authors:** Shabbir Ahmed, Fatema Akter Tanbi, Hossain Zillur Rahman

**Affiliations:** = Power and Participation research Centre (PPRC), Dhaka, Bangladesh

**Keywords:** Maternal mortality, Maternal death, 3 zeros agenda, last mile challenge, Bangladesh

## Abstract

**Background:** Reducing maternal mortality to zero is one of the 3 targeted agendas of the International Conference of Population and Development (ICPD). Though global improvements in align with SDGs commitment have witnessed comparative development but in the case of reaching zero agenda there are still much more improvements to make.

**Objectives:** Understanding the bottom-level reality with field experiences while also paying attention to the policy voices to understand the last mile stumbling blocks.

**Methods:** Reality check along with expert opinions working at the field level and beneficiary consultations were the primary components for this study. Three specific modalities (i) field-level consultations with the stakeholders, (ii) facility inspection and (iii) roundtable discussion were the specific tools used for this study.

**Results:** Findings show that in Bangladesh, substantial inequities exist both in the use of facility-based maternal healthcare with scarce and the absence of competent birth attendants for home births. Moreover, socio-cultural norms and the socioeconomic status of the families contribute to maternal deaths. The poor family setting, the hospital referral system, and the familial understanding of the overall maternal healthcare are largely responsible for maternal deaths.

**Conclusion:** All these issues related to maternal deaths need proper attention to leap forward to overcome the last mile challenges. Determined focus on defining and identifying the last mile population, breaking the pedagogical limitations and institutional barriers with an additional focus on the last mile research to reach the underserved population is imperative to reach the last mile.

**Problem:** Improving maternal health is one of three ICPD 3 zeroes priority, and a nested target in the third goal of the Sustainable Development Goals (SDGs). Alarmingly, maternal mortality is the third highest cause of death among women aged 15 to 49 in Bangladesh contributing to stalling the progress ahead in achieving SDGs.

**What is Already Known:** The business as usual and contextual factors including poverty, distance to health facilities, lack of information, inadequate services, cultural practices, education coupled with governance and macro-structural factors, private sector infrastructure investment and health finance are greatly responsible for maternal mortality to overcome last mile challenge.

**What this Paper Adds:** This study identifies barriers that limit the availability and access to quality maternal health services at all levels of the health system focusing the grass root maternal services. Underlines the prevailing inadequate and absence of services delivery and suggests necessary steps to address the last mile challenges towards ending preventable maternal deaths,

## Introduction

The high peak of maternal mortality is a conspicuous and dominant worrisome trend particularly in poorer nations. Developing nations bear the major share of this fatality, notably Southern Asia and Sub-Saharan Africa with two-thirds of the total maternal deaths (WHO, 2019b). A set of both direct and indirect causes manifests maternal mortality. Direct maternal deaths include those brought on by conditions such obstetric hemorrhage, hypertensive conditions, anesthesia difficulties, and cesarean section complications. While indirect causes are linked with “previous existing disease or disease that developed during pregnancy, and that were not due to direct obstetric causes but were aggravated by the physiologic effects of pregnancy”. In general, major triggering factors for the maternal deaths are severe bleeding, infections, pre-eclampsia and eclampsia, complication from pregnancy and childbirth and unsafe abortions (WHO, 2023). Though the improvements over the decades in reducing maternal deaths is remarkable with improved health systems targeted to implement interventions aimed at reducing social and structural barriers. Still in order to achieve the SDG target 3.1 and other related commitments systemic health failure at the grass root level is critical as inadequate access and poor quality of care for the marginalized women and girls disproportionately impedes the global goal (Govender, Topp, & Tuncalp, 2022).

Thus last mile challenges with disrupted systemic flow of the services in grass root communities and regions thwart initially set target. Poor structural design and outfitting of the services and interventions at the initial stage without considering the last mile issues hinder the progress. In such circumstances considering the bound of costs and time traditional approaches are subjected to reconsideration in the face of radical technological solutions for achieving the advantages of leapfrogging. With this in mind this paper investigates the issues and challenges in last mile maternal healthcare services and further ways in achieving the global target of reducing maternal mortality.

### Last mile challenges in maternal mortality

The radical development in maternal health stemmed from the Millennium Development Goals (MDGs) further enshrined and rightfully reflected in the Sustainable Development Goals (SDGs) target 3.1 to ensure and sustain the development achieved for further prominence. Target 3.1 explains that by 2030, the global maternal mortality ratio (MMR) will be reduced to less than 70 per 100,000 live births, and no country should have an MMR of more than 140 per 100,000 live births. This emerging call is in alignment with the International Conference on Population and Development (ICPD) 3 zeroes agenda, reducing maternal mortality as one of the 3 agendas, further unequivocally soliciting global urgency. In translating the commitments of both SDGs and ICPD into effective action for the last-mile population is a perceptible challenge. It is also critical to think about the persistent global health inequalities and the commitments on the population front and not to leave the population who were left behind during MDG periods. On top of that the widened chasms in global health inequality due to the COVID-19 pandemic within particular groups, who are living in poverty, and the reversed progress in health development and opportunities for the marginalized and sub-population groups are quite striking. As a result, all the barriers in service delivery have been reinforced as last-mile challenges during the phase of complacency.

Despite the bleak scenarios, many countries have made significant progress, but major gaps in the equitable coverage of crucial interventions for maternal and perinatal health continue to be barriers in many contexts. Globally maternal mortality is reduced by 38% from 2000-2017 (WHO, 2019a). Though maternal mortality is observed declining, the rate is not similar across the countries. Considering the current pace of development an estimated more than one million lives coupled with the burden on maternal health and well-being will fall short to achieve the global Sustainable Development Goals (SDG) target. Additionally, an estimated 810 women from low-and middle-income countries die every day due to complications of pregnancy and childbirth (WHO & UNFPA, 2021). It is perceptible that any of these global targets would not be able to make progress if the healthcare needs of all people down the last mile are rightfully addressed. In order to achieve the SDG targets, a human rights-based, comprehensive approach to sexual, reproductive, maternal, and newborn health is essential. This approach also forms the basis for a strengthened health system, which facilitates the successful implementation of universal health coverage. Besides, defining the last-mile populations and outlining their needs and expectations, and broadening the concerns in terms of how research best engages at or with the last mile is of utmost critical.

### Global and Bangladesh context of maternal mortality and last mile challenges

Global focus on reducing maternal mortality was the result of sequential initiatives since the 1980s that began with the Safe Motherhood movement in 1987 (Shiffman & Smith, 2007). Further, the robust accord was founded through the incorporation as one of the Millennium Development Goals. Subsequently, further emphasis was imposed upon realizing the importance of reducing maternal deaths in the long-term vision of Sustainable Development Goals retorting the global call for reducing maternal mortality to 70 per 100,000 live births by 2030. In order to achieve this ambitious targeted result, it is crucial to set informed and goal-oriented targets for reproductive health and up-to-date data tracking (Bhutta et al., 2010). It is also critical to be mindful of particular challenges like misclassification of maternal deaths with other vital registration and medical certification of deaths, sampling error due to recall bias as less maternal death is reported, large non-sampling error with repeated and overlapping measurements in the survey and census, various estimation in the demographic calculation of reproductive-age mortality from all causes, and absence of specific model in case of sparse and limited data (Leone, 2013) (Helleringer et al., 2013) (Cross, Bell, & Graham, 2010). Nevertheless, despite long-term attempts to reduce maternal mortality by offering antenatal care, emergency obstetric treatment, and skilled attendance during delivery, the risk of dying during pregnancy or childbirth is still quite high for women in developing countries (Campbell & Graham, 2006). Data between 1990 to 2005 shows a decline in maternal mortality from 179 per 100,000 live births to 132 per 100,000 live births across Latin America and the Caribbean whereas across Asia it has decreased from 410 to 329 per 100 000 live births. Nonetheless, in sub-Saharan Africa, the MMR was quite high, at more than 900 per 100,000 live births (Campbell & Graham, 2006). Considering the current progress to rein maternal deaths it is critical to design effective interventions considering last-mile challenges. Evidence-based monitoring and evaluations of these interventions are also essential to determine the efficacy and further modifications. This particular setting is too compatible with developing countries where the rate of maternal deaths is highest and the services delivery is too incompetent.

In Bangladesh, based on examining policy formulation and programme initiatives, access to maternal and reproductive healthcare services, and relevant contextual factors, it is concluded that related factors (education, access to information, and socio-economic conditions) directly connected to healthcare services and services distal such as accessibility (transportation and mobile phone) are influential in determining maternal health. It has also found that girls with at least some secondary education believe themselves to be empowered and have the ability for making responses to maternal complications situations and are competent in navigating healthcare services (El Arifeen et al., 2014). This clarifies the context that in order to achieve the predetermined targets of maternal mortality there is a pressing need to go over and beyond the healthcare system. However, health factors including postpartum hemorrhage, eclampsia, and abortion-related complications need proper attention in terms of available services delivery. Though the National Strategy for Maternal Health 2019 to 2030 devises the operation plan under the Health Nutrition and Population Sector Development Programme (HPNSDP) to provide Maternal and Neonatal Health (MNH) services with Basic Emergency Obstetrics and Newborn Care (BEmONC) and Comprehensive Emergency Obstetrics and Newborn Care (CEmONC). Additional focuses, in order to fully incorporate the continuum of maternal health care services, on the issue of maternal morbidity, family planning, menstrual regulation, and nutrition have been equally planned (MoHFW, 2019). But the question of efficacy in providing desired services in all of these domains is still questionable in terms of equipped facilities and staff from primary to tertiary level. Moreover, in terms of catering desired services in maternal care, the quality gap is paramount in the form of long waiting hours, facility cleanliness, terse consultation period, negligible and disrespectful care, paucity of essential drugs, and unexpectedly costly services (BBS & UNICEF, 2014). Moreover, privatization of the healthcare services pushes inappropriate diagnostics and treatments which are yet to be addressed properly by the government. Additionally, a significant cumbersome challenge of adolescent pregnancy as a result of child marriage lights on another critical issue which is one-fifth in the country as 59% of adolescents get married by the age of 18 and 21% by the 15. Data also reveals that 41.4% of women in Bangladesh give birth at the age of 18 and almost 58% at the age of 20 (MoHFW, 2006).

Moreover, out-of-focus policy priorities entail the reality of unreached adolescents which manifests in three critical areas - Sexual and Reproductive Health (SRH), human capital enhancement, and civic education. Breaking the barriers of limited pedagogic and mindsets issues is critical in this regard in terms of elaborated Sexual and Reproductive Health Rights (SRHR). It is also critical to broaden the focus lens equally to rural areas and consider the urban poor for catering better family planning and reproductive healthcare services.

### Why Maternal Mortality is a priority?

It is noteworthy how Bangladesh’s recent progress on health indicators has put the country on pace to meet its MDG targets, notably the MDG 4 on child survival. However, despite a projected annual rate of 5.5% and a 66% decrease in maternal mortality over the past two decades, Sen et al. (2018) Bangladesh is perplexed by the paradox that the recent progress on health indicators is not sufficient to overcome the last-mile challenges. According to reports, from 2015 to 2017, 37.2% of expectant women obtained the four required antenatal visits from qualified healthcare professionals. In 2020, this proportion was raised to 50%. Between 2015 and 2020, the percentage of pregnant women receiving postnatal care within 48 hours from a caregiver with medical training grew from 36% to 50% (MoHFW, 2019). However, the rates are still insufficient as it is anticipated to rise to 100% by 2030.

Although the Maternal Mortality Ratio (MMR) from 434 per 100,000 live births in 2000 has further reduced to 173 in 2017 (WHO, 2019b), whereas the SDG target of MMR is less than 70 per 100,000 live births. Thus, the current rate is regarded as unacceptably high, with almost 20,000 Bangladeshi mothers passing away each year from pregnancy and childbirth-related causes (Mitra, Al-Sabir, & Anne, 1997). This is one of the last mile challenges which is dependent not only on the medical system and women’s health, but also on human behavior and socio-cultural norms. The poor maternal health situation in Bangladesh is caused by a variety of significant socioeconomic and health system concerns. Currently, 14.7% of infants are born in medical facilities, 7.6% in private hospitals or clinics, and 7.1% in government institutions (Kamal, 2013). This low percentage of institutional medical care for deliveries and complications is undoubtedly one of the main underlying causes of Bangladesh’s poor maternal outcomes. According to recent research, iron deficiency anemia is common in Bangladeshi girls, especially in rural regions, and affects 50–90% of expectant mothers in the country (Rahman, Parkhurst, & Normand, 2003). Pregnancy anemia contributes to intrauterine growth retardation, resulting in low-birth-weight kids, and can increase the chance of maternal death. Furthermore, malnutrition in mothers and adolescent girls imposes significant expenditures on the healthcare system through increased morbidity and early birth. The stagnant improvement of Bangladesh’s maternal mortality status is greatly influenced by all of these variables.

As a result, there is a need to address the issue of maternal care availability and utilization in Bangladesh, where several overlapping determinants affect women’s maternal health. There appears to be some evidence that advances in healthcare quality may enhance service utilization Thaddeus and Maine (1994), but other research has highlighted the socio-cultural barriers that can limit women’s use of professional treatment regardless of facility quality. Over the previous decade, Bangladesh has made significant progress in terms of health. However, comparable progress in maternal health has not been made. Maternal mortality is a measure of not only women’s health but also of the country’s health sector’s access, quality, and efficacy from the standpoint of health systems. While there is a general global consensus on the measures required to reduce maternal mortality, there are still a number of important information gaps in Bangladesh on how to successfully implement and employ those interventions. This is why there is a pressing need to overcome the last-mile challenges in maternal mortality to make significant progress in the health sector.

Understanding the commitments to improve maternal health stressed back to many more ambitious goals and summits, it is crucial to keep focused on the objectives and understands the critical obstacles in achieving the full-fledged targets. Considering the critical aspects of reaching last-mile challenges, in this research, we are keen to understand the last-mile challenges in maternal mortality by dissecting multi-stakeholders’ perspectives and finding possible solutions and strategies to leap forward in meeting the zero preventable maternal mortality target.

### Methodology

#### Study design and settings

In this research the semi-structured questionnaire/checklist were used in three broad different modalities to dive deep into the subject matters in achieving the study objectives and insights. The three main modalities were (i) field-level consultations with the stakeholders (both demand and supply side stakeholders), (ii) Facility inspection (iii) Roundtable discussion (inclusion of policy voices). Following these three modalities, we initiated a series of conversations and consultations and engaged policymakers, experts, grassroots/field-level workers, and participants formatted in **Figure 1**. The conversation was arranged in both a roundtable format and group facilitation. First, we initiated our activities with the consultations, one with the local level service providers and the second with the service receivers. Separate group consultations were arranged for understanding the existing service delivery architecture and gaps in reaching last-mile population. Secondly, facility inspection was targeted to understand how maternal healthcare services are catered to the beneficiaries. In this regard, the union health and family planning welfare center was visited to inspect the service delivery architecture modality. A critically significant reason for facility inspection has also covered the intention to scrutinize how the labor room or delivery room is designed and how far the ambient created in the facility encourages or reflects the needs and the demands of the pregnant women. Finally, a roundtable discussion was arranged to reflect the combined perspectives form diverse stakeholders. In this regard, policy makers and experts working in the field level catering maternal health services including healthcare professionals were gathered at the roundtable for both open and structured discussion. The roundtable discussion was moderated in a structured manner by the lead researcher leaving an auspicious space for unstructured discussions too.

**Figure 1:**
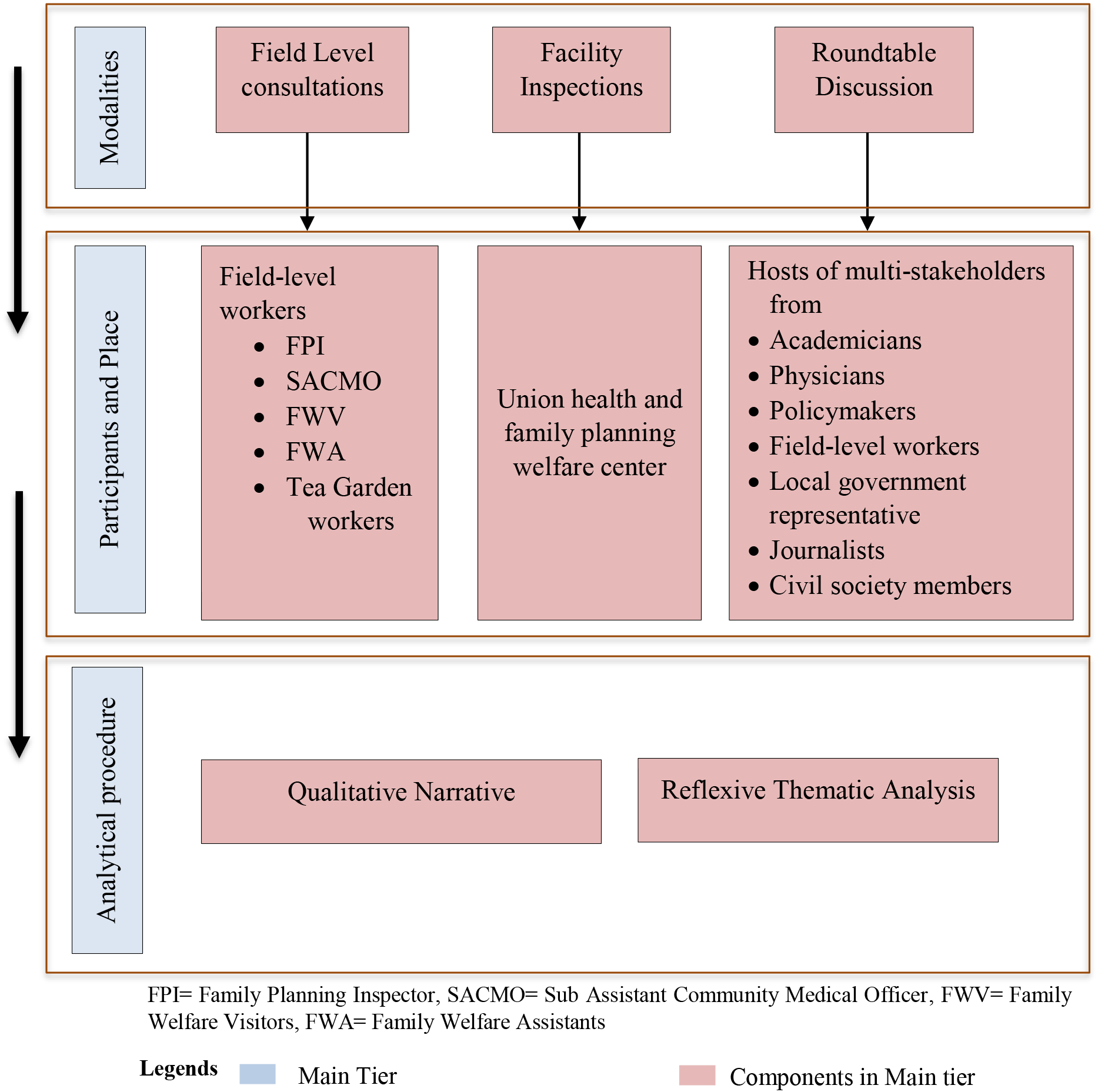
Methodological outline of the study in different tiers consisting of three modalities

#### Rationale for choosing three modalities

In this qualitative study, three modalities were selected (i) field-level consultations with the stakeholders, (ii) facility inspection (iii) roundtable discussion. The combination of these three modalities was auspicious in the case for understanding the situation of maternal mortality from diverse perspective (both supply and demand side reflections and shortcomings). Moreover, the combination of these three modalities was suited better to understanding the current situation and meeting the study objectives. Additionally, as a qualitative tool to reach the objectives and find out the targeted information these three modalities were best suited. The first modality provided necessary insights into understanding the root-level operations of maternal healthcare services from both the perspectives of service providers and service receivers to understand the challenges and limitations. The second modality was more of an investigation of the existing facility and observing how the services are in the physical condition and designed from the provider’s end.

This modality was crucial in terms of understanding how top-down designed architecture is in place to cater services to the bottom and how much the needs and aspirations of the bottom (pregnant women) are reflected in the architecture. The third modality was an efficient one in putting multi-stakeholders related to maternal mortality in a solid conversation in dissecting the last-mile issues of maternal mortality.

#### Data collection and reporting

Data collection procedures for the three modalities were some of a similar pattern across the study time period only the variations were in number of participants and time allocation. The time stamps for gathering information for all three modalities were different for separate modalities. The first modalities were 1 hour 30 minutes each to cover up the discussions. The second modality of facility inspection was consisted of 1 hour comprised of walk and talk investigation. The third modality was longer compared to the other two as this session was designed to discuss the whole range of issues and perspectives from different arrays of stakeholders. Though the reporting structure was the same for all modalities. The lead researchers led the conversations required for the three modalities and the other researchers took the preliminary notes and inspected the necessary details pertinent to the study. Each group discussion was audio-recorded and transcribed later for the convenience of clarity. In addition, designated researchers took field notes which were refined and synthesized relating to perceived dynamics and interactions between participants and merging study objectives. Additionally, to make the findings coherent the substantial considerations of the gray literature were also of the focus and qualitative narrative analysis was used in such instances.

#### Study site and analytical framework

**Figure 2:**
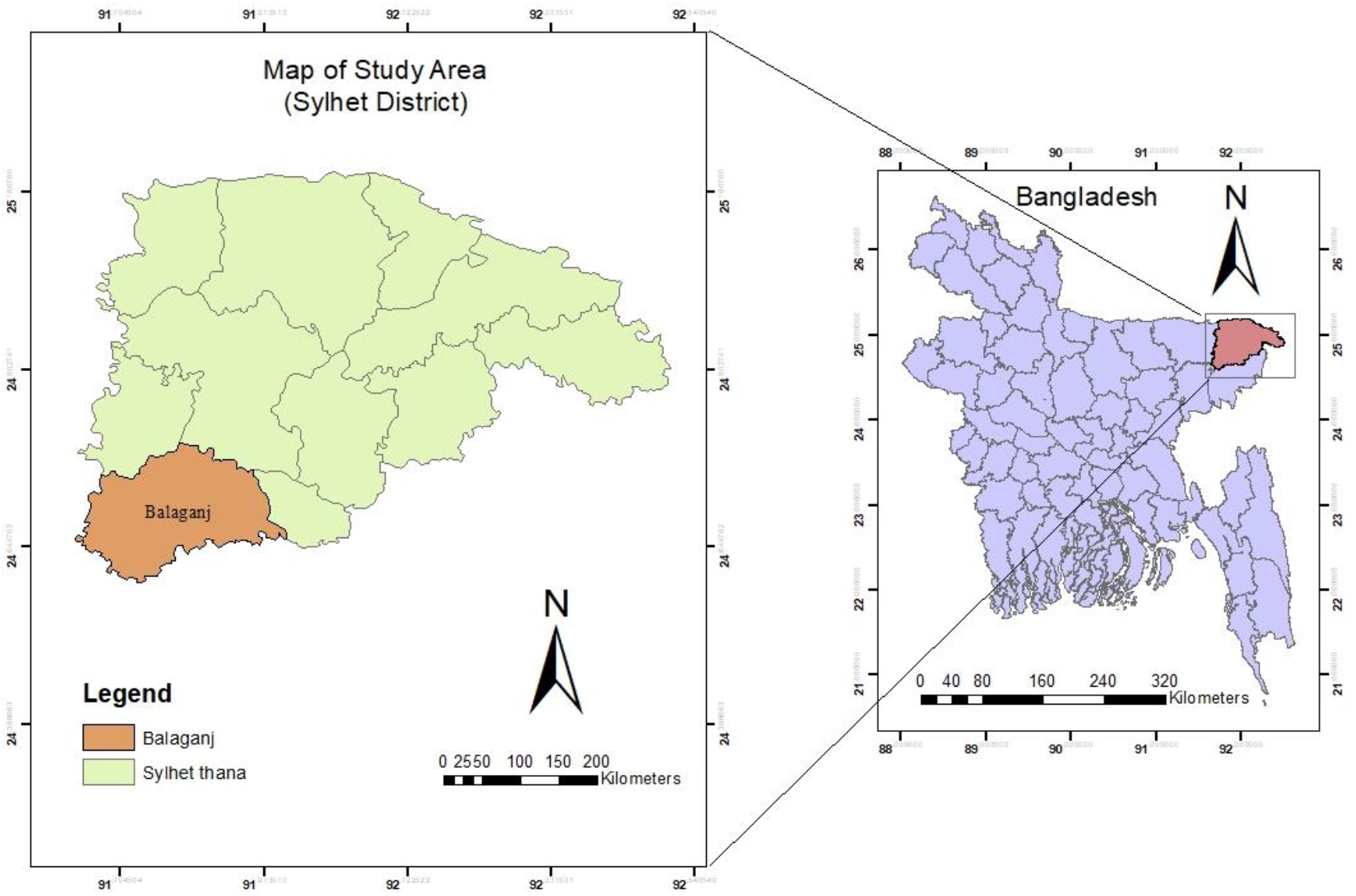
Study area

Maternal mortality is the third highest cause of death among women aged 15 to 49 in Bangladesh, accounting for 14% of all such deaths (El Arifeen et al., 2014). In our study, all three selected modalities were operated in Sylhet districts, which have the highest maternal mortality rate per 1,000 women and the fourth highest in maternal mortality per 100,000 live births in Bangladesh (NIPORT, ICDDRB, & MEASURE, 2019). Analytical framework of this study to present the findings in an organized manner the reflexive thematic analysis was followed Braun and Clarke (2006) which provided a background of theoretical flexibility in making active decisions leveraging the independent assumptions throughout the research process. Moreover, prioritizing the subjectivity of the research and as a research tool in the analysis process this method is one of the qualitative research paradigms as the process itself looks for patterns of shared meaning across the qualitative information. Most importantly, a frequently used tool in the analysis of clinical practice. Additionally, the epistemological underpinning of this analytical procedure is also of great relevance to this study. Furthermore, to present the findings in a coherent format thematic qualitative analysis was also used.

#### Ethical considerations

All data used in this study was collected with oral consent and analyzed anonymously. Confidentiality was ensured as required under EU regulation 2016/679, of 27th April, concerning the General Protection of Personal Data and according to the Organic Law 3/2018 for the Protection of Personal Data and the guarantee of digital rights. However, the institutional ethical approval for this research was opted for considering the purpose of education research with minimal risk to participants in terms of well-being, privacy, and confidentiality. Moreover, this research also aimed to understand the quality of services where the intent was to inspect the existing services architectures and find the gaps in services. Additionally, this research also focused on administering the anonymized survey with no identifiable information collected from the participants.

The subsequent chapter and sections are the findings of this study, which has been organized in such a manner to give a little preemptive to the study topics getting to know service delivery in a nutshell with the current situation.

## Findings

### Factsheets Services Delivery Architecture and Field Reflections

In Bangladesh, maternal healthcare services are catered to by both formal and informal sectors. Among the formal service sector non-public sector or private care, providers are widely available, and informal services are highly delivered by the Traditional Birth Attendance (TBA). Most notably in Bangladesh, a higher number of deliveries are performed by the TBA than institutional deliveries and C-section deliveries are dominant in private health facilities. Findings show that economic inequality forces women to avoid institutional deliveries ^21^. Despite the necessity, limited information and accessibility coupled with inadequate services and manpower stand as barriers to availing desired maternal healthcare facilities. There are two mainstream facility approaches through which maternal healthcare services are reached.

#### (i) Community-based approach

Family Welfare Assistants (FWAs) primarily carry out the operations of maternal health services at the community level through the distribution of family planning methods, such as oral pills and condoms, along with referrals for other methods. Besides, providing Antenatal Care (ANC) and referring to high-risk pregnancies also entail the duties of the FWAs. Catering maternal healthcare services from household to the sub-district level with the help of two separate ministries, the Ministry of Health Services and Family Planning, were the main objectives of community-based healthcare facilities. Other service provider agents such as Family Welfare Visitors (FWVs) provided the ANC, PostNatal Care (PNC), and basic Essential Obstetric Care (EOC) from the union-level Health and Family Welfare Centres (HFWCs). Describing the duties of grass root healthcare workers, one FWA from the first group consultation retorted:

> “At first, we go to the home of the expecting mother and register the necessary information. Then pregnancy counseling regarding Neonatal Care (NC is given which is at least 4 times during the pregnancy. The areas with limited transportation facilities are set to provide counseling services at least 1-2 times. Describing reasons for maternal mortality she said religious sentiment is partly responsible. By holding religious sentiments women are not willing to go outside of the home, even sometimes they hide the news of pregnancy. Many families think it is a matter of shame so they don’t want anyone else to know about it. The proportion of institutional delivery was 25, and house delivery was 20 out of her 45 pregnant attendants”

#### (ii) Facility-based approach

Facility-based approaches in maternal healthcare services were aimed to deliver services of Emergency Obstetric Care (EmOC) along with renovating and upgrading the existing service facilities with logistics and equipment, and human resource development (HRD).

In order to grasp a fuller picture of the service delivery architecture of maternal healthcare under the two distinct ministries of Health and Family Welfare (MoHFW) the following graphical illustration:

**Figure 3:**
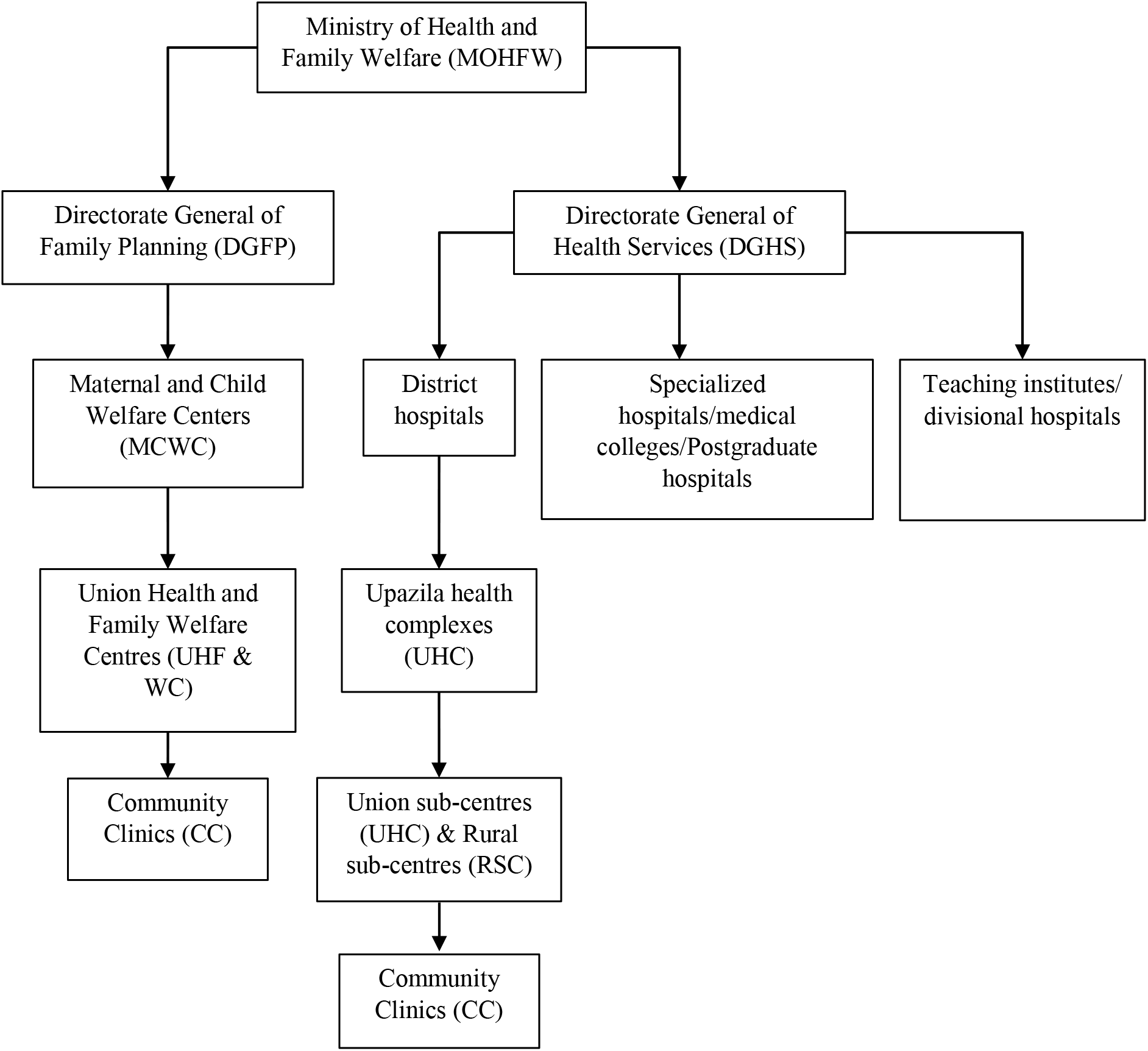
Maternal healthcare service delivery architecture in Bangladesh

### Health system architecture

The Ministry of Health and Family Welfare (MoHFW) works in two different wings to cater to maternal healthcare services. The first one is the Directorate General of Health Services (DGHS) and the second one is the Directorate General of Family Planning (DGFP).

DGHS implements all health programmes with the following architecture in three tiers of administrative coverage.

**Figure 4:**
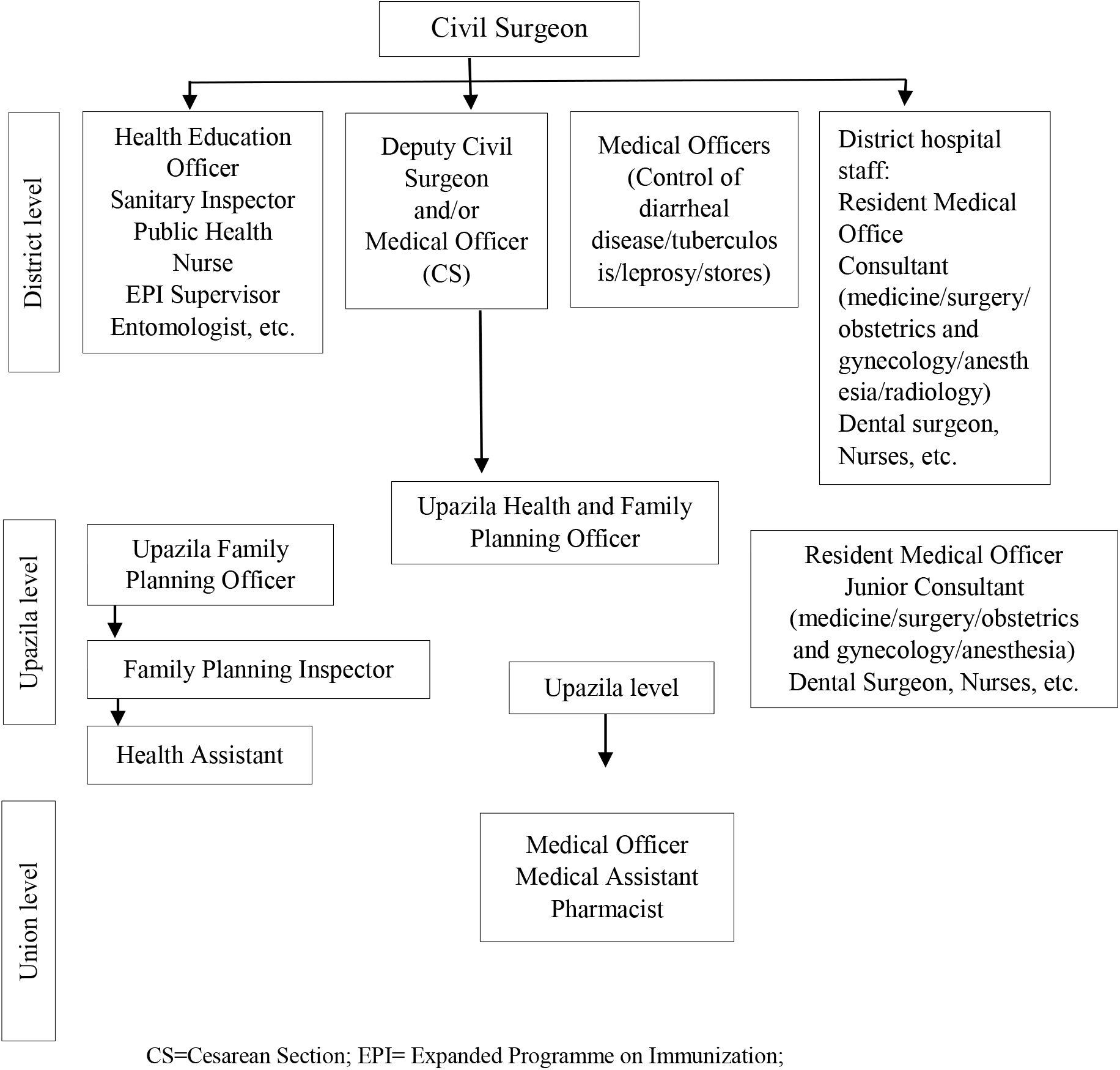
Health system architecture implemented by the Directorate General of Health Services (DGHS) at different administrative levels in Bangladesh, Source: (Perry, 2000)

On the other hand, according to Rahman et al. (2003) DGFP implements Family Planning (FP) programmes with the following structures

- 3275 Union Health and Family Welfare Centres (UHFWCs) exist to serve the 4,470 unions.
- There are Upazila Health Complexes with 31 beds in 391 rural Upazilas, 64 district hospitals,
- 13 Government medical college (MC) hospitals,
- 6 postgraduate hospitals and
- 25 specialized hospitals
- 54 Maternal and Child Welfare Centers (MCWCs)
- Community Clinic (CC) at the village level for every six thousand population
- The total number of registered doctors in the country is around 27,546
- There are 15,804 registered nurses
- There are 40,773 hospital beds, out of which 29,402 are located in government hospitals

**Figure 5:**
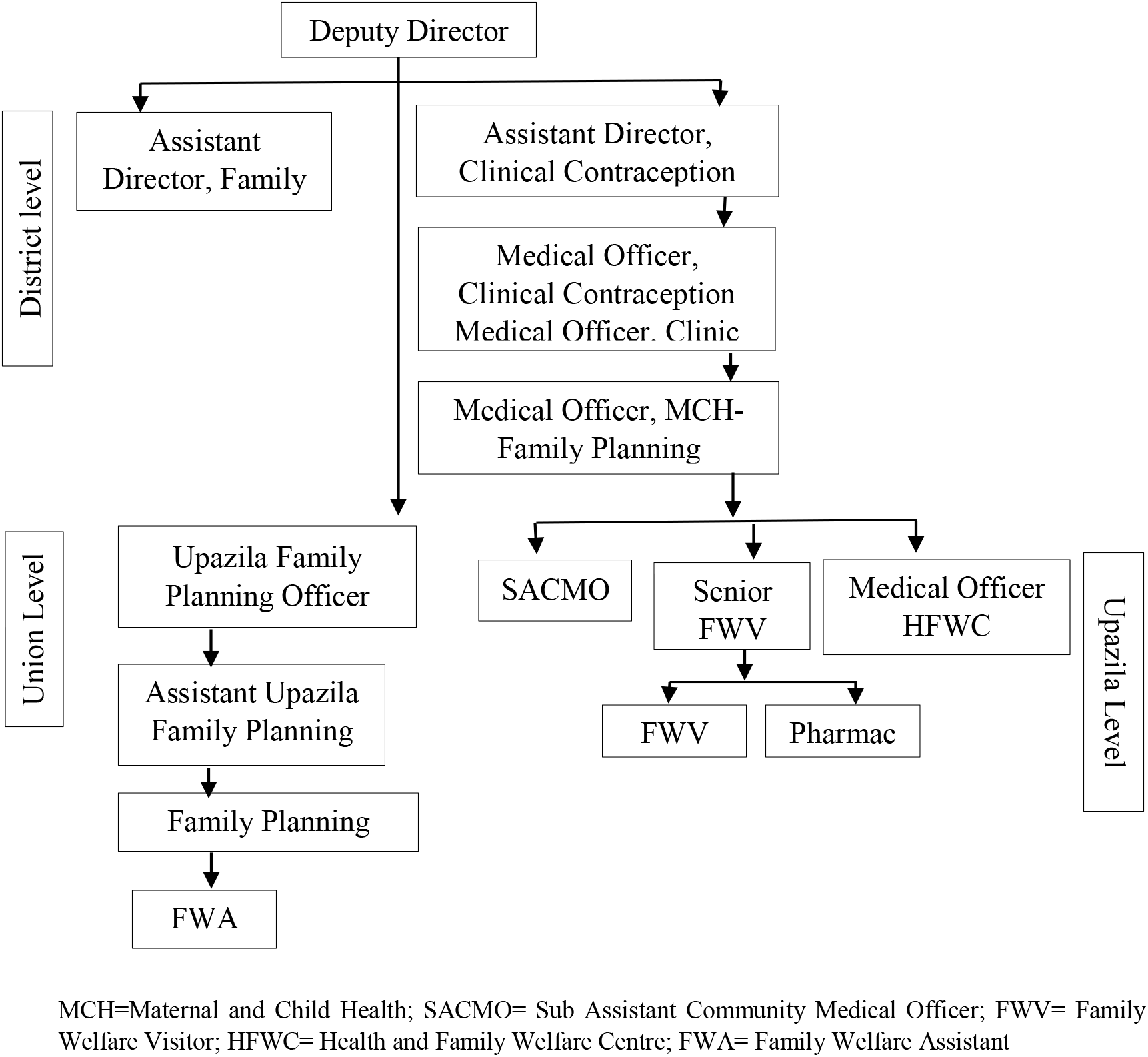
Health system architecture implemented by the Directorate General of Family Planning (DGFP) at different administrative levels in Bangladesh, Source: (Perry, 2000)

### Gaps in the service delivery architecture and Implementation challenges

#### Lack of community engagement

Though the service delivery architecture is stressed at the community level, there is a lack of community engagement and participation in maternal healthcare services, which limits the effectiveness of health interventions. While Antenatal Care (ANC), Postnatal Care (PNC), and Emergency Obstetric Care (EOC) are prioritized at the community level, many women still face barriers to accessing this quality care due to ineffective community participation. Reflecting on the opinion of the filed level community people one woman from the tea garden reported that

> “The rate of normal delivery at home with the help of a midwife is higher for the garden workers in our community as it is convenient financially unless further complications are faced.”During complicated pregnancy situations, there are reported cases of maternal deaths in the tea garden area due to the long-distance health facility services.

During our consultation an aged women sieve through her experiences and retorted,

> “In other remote tea gardens, including ours, there are witnessed cases of maternal deaths while reaching institutional facilities because of the long-distance issues of the healthcare facilities.”

#### Inadequate information dissemination

Many expectant mothers are not fully informed about their healthcare options limiting their ability to make informed decisions about their care. Though there are community-based programs, mass media campaigns, and training for healthcare providers to effectively communicate with patients. Without proper information and understanding of the delivery process, expectant mothers may not be fully prepared for childbirth, which can increase the risk of complications. Describing systems inefficiency one FWA said,

> “As we keep the online track of pregnant mothers in our respective working areas, there is a problem in updating the list of pregnant mothers and sometimes if delivery is conducted, it is not taken off the list and this increases confusion. There is also backlog issue and lack of training among the field level workers further worsen information automation problem using the online application.”

#### Limited access to quality care

The quality of maternal healthcare services is often poor, particularly in rural areas, which results in inadequate care for expectant mothers. The shortage of skilled healthcare professionals, transportation obstacles, and a lack of resources and equipment, particularly in rural areas, restrict the availability of quality maternal care. Reporting the limited access to care the FPI stated,

> “The financial issues of the families and transportation problems due to the remoteness of the area stand as the primary reasons for the family members not wanting institutional delivery and opt for home delivery even without skilled birth attendants.”

#### Inefficient healthcare delivery systems

The healthcare delivery system in Bangladesh is often fragmented, which results in inefficiencies and limits the availability of quality care. Many healthcare facilities lack the necessary equipment, medicines, and supplies to provide quality care, which can put expectant mothers and their babies at risk. Citing the inefficiency of the services FPI mentioned,

> “Every month there are demands for 18-20 deliveries. But the office time in the FWC is only open for 3 PM every day and there is no service for ANC in the centers. Moreover, manpower shortage makes it difficult to provide 24/7 services to pregnant mothers.”

#### Insufficient coordination between different stakeholders

There is often a lack of coordination between different stakeholders involved in maternal healthcare services, which can result in duplication of efforts and waste of resources. At the roundtable discussion, one expert excerpted

> “Inadequate services are provided by institutions, which results in maternal deaths caused by various factors such as insufficient medication, injections for eclampsia prevention. Our objective is to work together diligently to prevent any maternal deaths. Both named and unnamed organizations cannot provide optimal services due to various issues, including equipment problems. Therefore, everyone should be accountable in handling maternal healthcare.”

#### Weak governance structures

Weak governance structures in remote and rural areas limit the effectiveness of maternal healthcare programs and interventions in place to monitor and evaluate the performance of healthcare providers and ensure that they are delivering quality care. It has also limited required community engagement in maternal health services, with few opportunities for communities to participate in the design and implementation of maternal health programs. One expert from the roundtable discussion stated,

> “There is a need to strengthen the activities of the 40,000 community clinics in the country. The shortage of skilled personnel in public and private clinics and hospitals is causing disruptions in services, which can be addressed by increasing the workforce.”

#### Insufficient data collection and monitoring

There is a lack of systematic data collection and monitoring of maternal healthcare services, which even makes it difficult to assess the impact of interventions and identify areas for improvement. Furthermore, the result of weak health information systems is reflected in poor decision-making. Addressing data inaccuracy as an issue, a representative from the Department of Women’s Affairs cited,

> Even health experts and researchers do not receive accurate information, emphasizing the need for a long-term plan to reduce maternal mortality while ensuring the accuracy of information. It is crucial to be vocal about this issue and take further action to reduce maternal mortality.

### Discussion

Understanding the priority set in the global agenda including SDG and ICPD commitments it is critical to consider the maternal mortality issue with great gravity and find out the possible hurdles in the way of achieving the target. In this study, we took a deeper dive into the issue of maternal mortality dissecting the perspectives from different angles with a different methodological approach.

Findings from this study explain that maternal death is a complex issue and rested not only in the medical system and health status of women but also depends on human behavior, sociocultural norms, and socioeconomic status too. Structural factors such as language, social habits, religion, and intermediary factors including place of residence, childbearing mother’s age, parity, and women’s exposure to communication media regarding maternal health messages are influential in maternal mortality and catering to maternal healthcare services. Additionally, the influence of structural factors on intermediary factors was also critical in maternal health either independently or coupled with other structural factors (Hamal, Dieleman, De Brouwere, & de Cock B, 2020). In our study, we found structural factors such as religion plays a vital role in determining the willingness to access maternal healthcare services. By holding religious sentiments, particularly in Muslim families, women are unwilling to go outside the home, even though sometimes they hide the news of pregnancy. Many families think it is a matter of shame so they do not want anyone else to know about the news of pregnancy. Similar findings reported that compared to Hindu women Muslim women were 4 times less likely to visit for ANC care (Hamal et al., 2020). Evidence suggests that reproductive and maternal healthcare-seeking behavior is largely influenced by gender roles due to patriarchal societal structures.

This study also signifies the problem of inadequate and skilled healthcare workers at the field level. Timely referral systems and limited time-bound healthcare services at the local level also fail to cater the desired services to the mothers. Findings from a nationally representative study conclude that due to inadequate and timely referral mechanisms, women with dangerous complications of convulsions or excessive bleeding and other highly time-sensitive complications treatment are delayed six or more hours due blurry thin line in deciding the treatment, which is significant than the travel distance and delays in receiving treatment (Koenig et al., 2007). Equipped and full-fledged services provisions at the bottom-level services centers are also questionable. This study finds that despite having an established health center aimed at catering to maternal healthcare has been left unutilized due to the absence of a tube well. Such inequity overrides progress to sustain a steady pace across all segments of society in accessing and providing quality maternal health care services at the grass root level. This ultimately becomes a concern for maternal health outcomes and access to healthcare services for achieving broader goals inclusive of both SDGs and ICPD (Gwatkin, Bhuiya, & Victora, 2004).

Considering the challenges at the bottom level in catering to maternal healthcare services and ensuring progress toward global commitments last-mile challenges are critical to have full attention. Being informed from the work of Davison et al. (2021) in addressing the last-mile dilemma in the research can be also extended in the case of maternal mortality. The extension of the knowledge can further be stressed with the following framework in the maternal mortality last mile challenged areas.

### Leaping forward to Address Last-mile Challenges

In order to come up with a feasible and realistic framework for the maternal mortality last-mile challenges, it is critical to dissect the existing milieu of maternal health services and associated factors to comprehend and structuralize ways out of the last-mile challenges. In order to initiate the process, the following three basic questions need to be answered.

**Figure 6:**
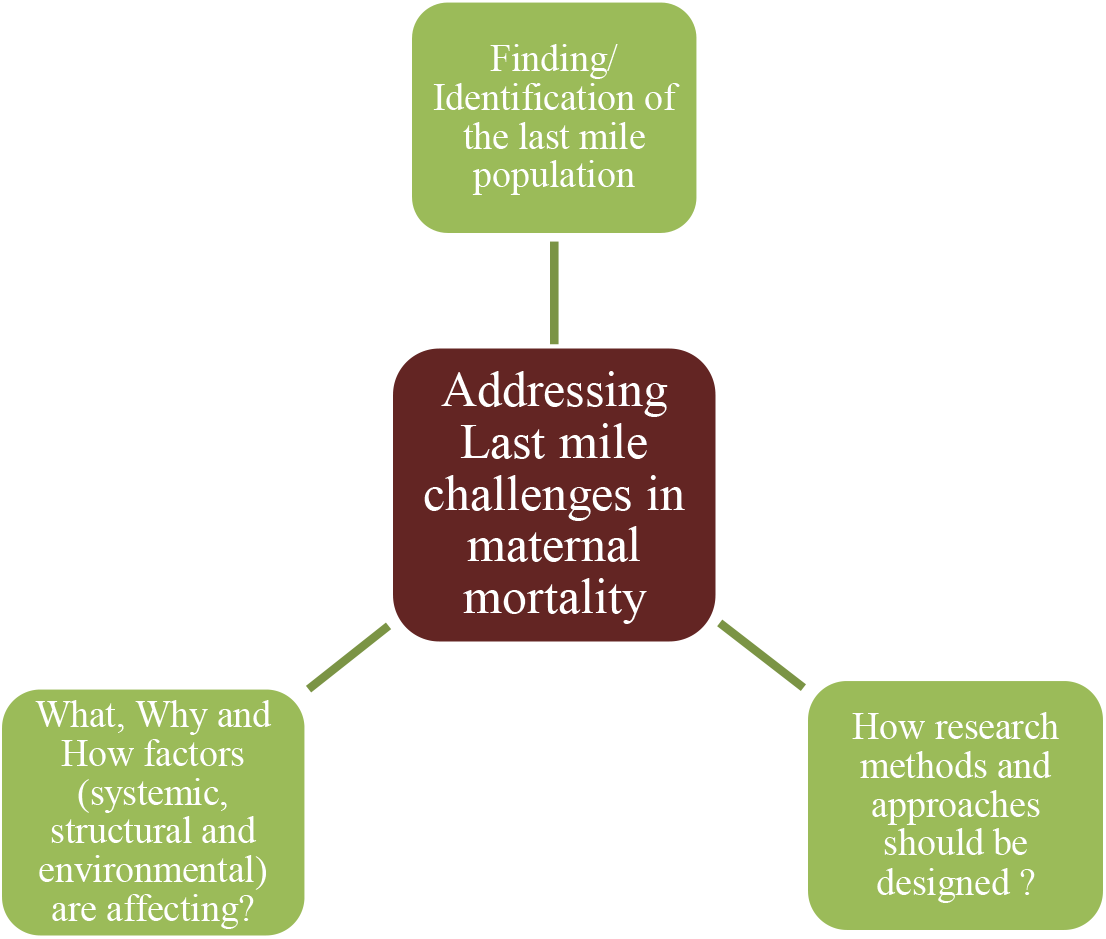
Leaping forward with the triangular perspectives

#### Identifying last mile population

As maternal mortality is not uniformly distributed ^28^ it is critical to find out and define people on the margin with poverty, disability, and other social ailments with geographically disadvantaged connectivity issues and need proper mapping and linking them with the existing services is one of the possible ways. Finding people without social and political voices and advocating for them mainstreaming their needs with equitable health services is critical in this perspective too. The availability of updated data on maternal mortality is also critical to differentiate the direct and indirect causes of maternal health and provide necessary interventions. This task of accurate data capture is difficult as many countries with the advanced statistical system have their own share of complicacies (WHO, 2005).

##### Box 1

###### How to Identify Last-mile Population?

- Rigorous needs assessment is critical to identify the communities and populations with the greatest need for maternal healthcare services, as well as the specific barriers that are emanating in accessing healthcare services.
- Full-fledged mapping of healthcare facilities and services to identify the communities that are farthest from care, as well as areas with limited access to quality maternal healthcare services require full investigation.
- Active and collaborative engagement with community leaders and stakeholders, such as local government officials, healthcare providers, and community-based organizations to identify the specific needs of the last mile population and the barriers they face in accessing maternal healthcare services.
- Periodical or continuous surveys and focus groups with expectant mothers and their families gaining valuable insights into the barriers they face in accessing maternal healthcare services, as well as the types of care and support that they need is a potential window.
- Innovative use of technologies to empower with data and information, such as geospatial mapping and mobile health technologies, is also critical to identify the last mile population and track their utilization of maternal healthcare services.

#### Understanding the ‘how’ part with better research

Reflexive thinking, empowering people and communities, building capacity and knowledge, and breaking pedagogical limitations will be better articulated and translated into action if the research is concentrated on the ‘how’ part concerning the last mile populations (Davison et al., 2021). In this regard operational research for assessing and designing health interventions and policies for better service delivery is tantamount. Engaging stakeholders across the field to understand the factors from the demand side of the services and formulation of strategies and service delivery models for the last-mile population.

##### Box 2

###### Better Understanding of Last-mile Population?

- Research endeavor is tantamount important to identify gaps in knowledge about maternal mortality in order to clarify the areas where further research is needed and where interventions needs to be targeted.
- Analytical insights from the collected data and, interventions are equivocally important to develop and address the root causes of maternal mortality.
- Evaluating multiple interventions ranging from improving access to healthcare, increasing the availability of skilled birth attendants, and improving nutrition and education for women is imperative to boost further knowledge.
- Methodical systematic monitoring and surveillance are important to track progress and to identify new or emerging challenges to ensure that the problem of maternal mortality is effectively addressed over the long term.

#### Identifying the factors

Systemic, structural, and environmental determinants of health or of being among last mile populations also merits the in-depth investigation in mitigating last mile challenges. Intensive clarification of upstream and downstream factors inclusive of the systemic biases and processes that contribute to individuals or groups being ‘last mile is also instrumental (Davison et al., 2021).

##### Box 3

###### Governance and management of the system

In order to efficiently mechanize and run the service delivery architecture it is critical to have rhythmic coordination among the national to community level programmes of both the Directorate General of Health Services (DGHS) and Directorate General of Family Planning (DGFP) accompanied by the related departments. Duplication and overlapping of efforts and services in catering to maternal healthcare needs to be avoided with real-time data, information platform sharing and integration. Additionally, ensuring the fullest utilization of the resources is critically dependent not only on the establishment of facilities but on being equipped with the necessary tools and components. Part of the whole service architecture would be paralyzed even if all the major components are in place but a simple tool is missing.

###### Decentralized planning with a logistical supply chain

In order to reflect the effective community and local needs it is crucial to implement District Evidence-based Planning and Budgeting (DEPB) to strengthen district and upazila health system responses for inclusive programming and resource allocation curbing the equity gap among different social, economic, geographic, and ethnic strata. At the same time, it is also instrumental to effectively and efficiently plan the distribution time, curb systemic inefficiency, establish accountability, and lastly regular monitoring of the inventories. This way, local communities and stakeholders can provide regular feedback on the performance of maternal health programs. Improved coordination between procurement and distribution systems at all levels of service delivery needs additional focus with target-oriented attitudes.

## Conclusion

In Bangladesh, maternal healthcare services at all levels of the health systems need to recognize and address these last-mile challenges to cater services to pregnant women, their partners, and their families receiving high-quality medical care living in underprivileged communities. Based on the discussion of this paper, reaching the zero preventable maternal mortality goal is achievable with not only strong commitments but with concentrated attention focusing last-mile population, wide and active stakeholder engagement, financial commitments, and political willpower. The progress made along the way should not get stuck or lowered by the widening inequities related to gender, race, religion, and other socioeconomic characteristics moreover the looming setback shadow of Covid-19. It is also critical to understand and solve barriers to equitable health services, emphasizing the inclusion and priorities of the last-mile population. Moreover, broad rigorous encompassing of the other interrelated issues and approaches like sexual and reproductive health, family planning, nutrition, and gender-based violence also need equal treatment to progress ahead. It is also essential to keep in mind that, progress in addressing last-mile challenges in maternal mortality is highly dependent on robust health systems ensuring the high coverage of midwifery services with timely and competent healthcare facilities at all levels. Improving access to skilled birth attendance at delivery and institutional services availability are also critical components. Additionally, facilitating the oversight of governments’ responsibility in reducing maternal death with appropriate interventions for the most marginalized groups, such as rural populations and the impoverished is also essential. In the end, improving access to skilled attendance at delivery or health facility is not going to solve the maternal mortality problem at the last mile, mindful attention to the social-economic indicators also demands the same attention.

## Data availability statement

The data that support the findings of this study are available on request from the corresponding author.

## Ethical Approval

All participants gave written and oral informed consent for the study prior to undertaking the interviews and discussions. All participants were asked and consented to the interviews being audio recorded.

## Conflict of interest

The authors declare no conflicts of interest.

## Abbreviations

ANC: Antenatal Care
DGFP: Directorate General of Family Planning
EU: European Union
FWA: Field Work Assistant
FWV: Family Welfare Visitor
HFWC: Health and Family Welfare Centres
HPNSDP: Health Nutrition and Population Sector Development Programme
ICPD: International Conference on Population and Development
MDG: Millennium Development Goal
SRHR: Sexual and Reproductive Health Rights
MMR: Maternal Mortality Rate
MCH: Maternal and Child Health
PNC: Postnatal Care
SDG: Sustainable Development Goals
SACMO: Sub Assistant Community Medical Officer
TBA: Traditional Birth Attendant
MNH: Maternal and Neonatal Health
SRH: Sexual and Reproductive Health

## Data Availability

All data produced in the present study are available upon reasonable request to the authors

## Notes

### Competing Interest Statement

The authors have declared no competing interest.

### Funding Statement

This study did not receive any funding

## References

BBS, & UNICEF. (2014). Bangladesh Multiple Indicator Cluster Survey 2012-2013, ProgotirPathey: Final Report. Retrieved from https://www.unicef.org/bangladesh/media/1021/file/Mics2013.pdf

Bhutta, Z. A., Chopra, M., Axelson, H., Berman, P., Boerma, T., Bryce, J., de Francisco, A. (2010). Countdown to 2015 decade report (2000–10): taking stock of maternal, newborn, and child survival. The lancet, 375(9730), 2032–2044. 10.1016/S0140-6736(10)60678-2

Braun, V., & Clarke, V. (2006). Using thematic analysis in psychology. Qualitative Research in Psychology, 3(2), 77–101. doi:10.1191/1478088706qp063oa

Campbell, O. M., & Graham, W. J. (2006). Strategies for reducing maternal mortality: getting on with what works. Lancet, 368(9543), 1284–1299. doi:10.1016/S0140-6736(06)69381-1

Cross, S., Bell, J. S., & Graham, W. J. (2010). What you count is what you target: the implications of maternal death classification for tracking progress towards reducing maternal mortality in developing countries. Bull World Health Organ, 88(2), 147–153. doi:10.2471/BLT.09.063537

Davison, C. M., Bartels, S. A., Purkey, E., Neely, A. H., Bisung, E., Collier, A., Adams, L. V. (2021). Last mile research: a conceptual map. Global Health Action, 14(1), 1893026. doi:10.1080/16549716.2021.1893026

El Arifeen, S., Hill, K., Ahsan, K. Z., Jamil, K., Nahar, Q., & Streatfield, P. K. (2014). Maternal mortality in Bangladesh: a Countdown to 2015 country case study. Lancet, 384(9951), 1366–1374. doi:10.1016/S0140-6736(14)60955-7

Govender, V., Topp, S. M., & Tuncalp, O. (2022). Rethinking trust in the context of mistreatment of women during childbirth: a neglected focus. BMJ Glob Health, 7(5). doi:10.1136/bmjgh-2022-009490

Gwatkin, D. R., Bhuiya, A., & Victora, C. G. (2004). Making health systems more equitable. Lancet, 364(9441), 1273–1280. doi:10.1016/S0140-6736(04)17145-6

Hamal, M., Dieleman, M., De Brouwere, V., &, & de Cock B, T. (2020). Social determinants of maternal health: a scoping review of factors influencing maternal mortality and maternal health service use in India. Public Health Rev, 41, 13. doi:10.1186/s40985-020-00125-6

Helleringer, S., Duthe, G., Kante, A. M., Andro, A., Sokhna, C., Trape, J. F., & Pison, G. (2013). Misclassification of pregnancy-related deaths in adult mortality surveys: case study in Senegal. Trop Med Int Health, 18(1), 27–34. doi:10.1111/tmi.12012

Kamal, M. S. (2013). Preference for Institutional Delivery and Caesarean Sections in Bangladesh. *Journal of Health*, Population and Nutrition, 31(1), 96–109. doi:10.3329/jhpn.v31i1.14754

Koenig, M. A., Jamil, K., Streatfield, P. K., Saha, T., Al-Sabir, A., Arifeen, S. E., Haque, Y. (2007). Maternal Health and Care-Seeking Behavior in Bangladesh: Findings from a National Survey. International Family Planning Perspectives, 33(2), 75–82. Retrieved from http://www.jstor.org/stable/30039206

Leone, T. (2013). Measuring Differential Maternal Mortality Using Census Data in Developing Countries. Population, Space and Place, 20(7), 581–591. doi:10.1002/psp.1802

Mitra, S. N., Al-Sabir, A., & Anne, R. (1997). Bangladesh demographic and health survey 1996-1997. National Institute of Population Research and Training.

MoHFW. (2006). BANGLADESH ADOLESCENT REPRODUCTIVE HEALTH STRATEGY. Ministry of Health and Family Welfare

MoHFW. (2019). Bangladesh National Strategy for Maternal Health 2019-2030. Retrieved from http://dgnm.portal.gov.bd/sites/default/files/files/dgnm.portal.gov.bd/page/18c15f9c_9267_44a7_ad2b_65affc9d43b3/2021-06-24-11-27-702ae9eea176d87572b7dbbf566e9262.pdf

Niport, N. I. o. P. R. a. T., Icddrb, I. C. f. D. D. R. B., & Measure, M. E. (2019). Bangladesh Maternal Mortality and Health Care Survey 2016: Final Report. Retrieved from Dhaka, Bangladesh:

Perry, H. B. (2000). Health for all in Bangladesh: lessons in primary health care for the twenty-first century: University Press.

Rahman, S. A., Parkhurst, J. O., & Normand, C. (2003). Maternal Health Review, Bangladesh.

Sen, L. C., Ghosh, S., Huda, T. M., Mali, S. K., Touhiduzaman, A. S. M., & Akter, N. (2018). Knowledge and Skill of Traditional Birth Attendants in Hizla Upazila. International Joural of Innovative Research, 3(2), 44–51.

Shiffman, J., & Smith, S. (2007). Generation of political priority for global health initiatives: a framework and case study of maternal mortality. Lancet, 370(9595), 1370–1379. doi:10.1016/S0140-6736(07)61579-7

Thaddeus, S., & Maine, D. (1994). Too far to walk: Maternal mortality in context. Social Science & Medicine, 38(8), 1091–1110. 10.1016/0277-9536(94)90226-7

WHO. (2005). Health and the Millenium Development Goals.

WHO. (2019a). Executive summary: Trends in maternal mortality the United Nations Population Division. Retrieved from

WHO. (2019b). Trends in maternal mortality 2000 to 2017: estimates by WHO, UNICEF, UNFPA, World Bank Group and the United Nations Population Division. Retrieved from Geneva:

WHO. (2023). Trends in maternal mortality 2000 to 2020: estimates by WHO, UNICEF, UNFPA, World Bank Group and UNDESA/Population Division. Retrieved from Geneva:

WHO, & UNFPA. (2021). Ending Preventable Maternal Mortality (EPMM) A RENEWED FOCUS FOR IMPROVING MATERNAL AND NEWBORN HEALTH AND WELLBEING. Retrieved from https://www.who.int/reproductivehealth/publications/maternal-mortality-2000-2017

